# Hippocampal Avoidance Whole-brain Radiotherapy in Preservation of Neurocognitive Function for Brain Metastases: A Phase II Blinded Randomized Trial

**DOI:** 10.1101/2020.04.29.20080234

**Authors:** Wen-Chi Yang, Ya-Fang Chen, Chi-Cheng Yang, Pei-Fang Wu, Hsing-Min Chan, Jenny Ling-Yu Chen, Guann-Yiing Chen, Jason Chia-Hsien Cheng, Sung-Hsin Kuo, Feng-Ming Hsu

## Abstract

**Background:** Hippocampal avoidance whole-brain radiotherapy (HA-WBRT) shows potential for neurocognitive preservation. This study aimed to evaluate whether HA-WBRT or conformal WBRT is better for preserving neurocognitive function.

**Methods:** This single-blinded randomized phase II trial enrolled patients with brain metastases and randomly assigned to receive HA-WBRT or conformal WBRT. Primary end point is the decline of Hopkins Verbal Learning Test–Revised (HVLT-R) Delayed Recall at 4 months after treatment. Neurocognitive function tests were analyzed with a mixed effect model. Brain progression free survival (BPFS) and overall survival (OS) were estimated using the Kaplan–Meier method.

**Results:** Patients were enrolled from March 2015 to December 2018 with a median follow-up of 12.4 months. A total of 70 patients were randomized. No differences in baseline neurocognitive function existed between the two arms. There were no differences in any neurocognitive assessments at four months. At six months, patients receiving HA-WBRT showed favorable perpetuation of HVLT-R total recall (mean difference = 2.60, *p* = 0.079) and significantly better preservation of the HVLT-R recognition-discrimination index (mean difference = 1.78, *p* = 0.019) and memory score (mean difference = 4.38, *p* = 0.020) compared with patients undergoing conformal WBRT. There were no differences in TMT part A, part B, or the COWA test between the two arms at any time point. There were no differences in BPFS or OS between arms as well.

**Conclusions:** Patients receiving HA-WBRT without Memantine showed better preservation in late verbal memory, but not in verbal fluency or executive function.

## Introduction

Brain metastases are the most common brain tumors in adults. A total of 10%– 30% of cancer patients develop brain metastases during the course of their illness[1]. Its incidence continues to increase with advances in diagnostic modalities, effective systemic therapies, and improved survival of cancer patients[1, 2].

Historically, brain metastasis patients show poor survival, with a median of one month if left untreated and 3–6 months[3, 4] after treatment. Over several decades, whole brain radiotherapy (WBRT) has become the standard of care for treating brain metastasis[5], with an estimated response rate of 27%–56%[6, 7]. Several prognostic models have been developed to predict clinical outcomes. The recursive partitioning analysis (RPA)[8] proposed by the Radiation Therapy Oncology Group (RTOG) has been the most widely used. Recently, the Graded Prognostic Assessment (GPA) became the most common index for assessing brain metastasis outcomes[9]. Patients with a good prognostic score from either index have a predicted overall survival of around one year. Some specific subgroups of patients, such as oncogenes-driven lung cancer or luminal-A breast cancer, can show an estimated survival of around two years[10].

Despite improved survival in certain patients with brain metastasis, the toxicity of brain irradiation raises concerns. Late effects of WBRT generally appear ≥ 3 months after irradiation and could be irreversible and progressive; it is considered secondary to vascular injury, demyelination, and ultimately necrosis[11]. Symptoms range from mild lassitude to progressive memory loss and dementia[12-14]. The hippocampus is a known neurogenesis region[15] in adults, and brain irradiation causes hippocampal dysfunction, which results in memory defects and depression-like behavior[16].

Radiation dose and volume to the hippocampus can correlate with memory deficit[17]. Improved novel radiotherapy techniques make it possible to avoid the hippocampus while treating the entire brain[18, 19].

RTOG 0933 is a single-arm prospective phase II trial to evaluate the effect of hippocampus avoidance WBRT (HA-WBRT) on neurocognitive function[20]. It demonstrated the potential value in neurocognitive preservation of HA-WBRT using the Hopkins Verbal Learning Test–Revised (HVLT-R) tests. The HVLT-R-delayed recall decline at four months from baseline was reduced to 7% compared with 30% decline in historical control[21]. Here, we present a single-blinded, phase II randomized trial to compare neurocognitive function outcomes in patients with brain metastasis treated by either HA-WBRT or conformal WBRT.

## Methods

This study was approved by the institutional research ethics committee and was independently monitored by the institutional clinical trial center. Informed consent was obtained from each patient in written form. This randomized trial is registered in ClinicalTrials.gov with identifier (NCT number): NCT02393131.

### Study design and participants

Patients with histologically-proved non-hematological malignancy and radiographic evidence of brain metastasis outside a 5 mm margin around either hippocampus on gadolinium contrast-enhanced MRI obtained within 30 days prior to registration were eligible. Eligibility criteria included age 20 years or older, Karnofsky Performance Status ≥ 60, and life expectancy of at least 4 months. Patients with the following conditions were excluded: prior brain radiotherapy or radiosurgery to > 5 intracranial metastatic lesions or a biological-equivalent dose in 2-Gy fractions > 7.3 Gy to 40% of the volume of bilateral hippocampus from prior radiosurgery^[22]^. Other exclusion criteria included serum creatinine > 2.0 mg/dL within 30 days prior to registration, leptomeningeal seeding, contraindication to MR imaging, and severe active comorbidities as judged by investigators. Patients were not allowed to receive investigational systemic therapy during WBRT. Patients who met all eligibility criteria were randomly assigned to receive either hippocampus avoidance WBRT (HA-WBRT) or conformal WBRT.

### Treatments

The WBRT treatment was 3 Gy per fraction once per workday for continuous workdays (Monday to Friday) every week for 10 days, to a total dose of 30 Gy. A non-contrast treatment-planning CT scan of the entire head region was required to define planning target volumes and hippocampal avoidance zones. The pre-treatment brain MRI was fused semi-automatically with treatment-planning CT for hippocampal contouring. Contouring was carried out in accordance with the Radiation Therapy Oncology Group (RTOG) atlas based on RTOG 0933[20] with the assistance of experienced neuroradiologists. Treatment plans were designed with volumetric modulated arc therapy (VMAT or RapidArc) using 6MV photon for both HA-WBRT and conformal WBRT arms. Dosimetry criteria for radiotherapy planning are described in the **Supplementary Material**. Treatment was delivered with daily image guidance using online cone beam CT for 3D corrections.

### Assessment

All patients were evaluated at entry, during treatment, and after treatment at 1, 2, 4, and 6 months, then every 3 months until death or brain progression. Adverse events were graded according to the National Cancer Institute’s common toxicity criteria (CTCAE), version 4.

Neurocognitive function tests, self-reported neurocognitive function, and self-reported health-related quality of life were assessed at base line, at 1, 2, 4, 6 months, followed by every 3 months up to 24 months after WBRT unless brain progression or death occurred. The neuropsychological test battery included tests of memory, processing speed, executive function, and verbal fluency. Hopkins Verbal Learning Test-Revised (HVLT-R), Trail Making Test Part A (TMT-A), Trail Making Test Part B (TMT-B), and Controlled Oral Word Association (COWA) were conducted by blinded independent health professionals and data were recorded as raw scores, time, and word counts without normalization. Self-reported cognitive outcomes were assessed using the EORTC QLQ-C30, and FACT-Cog version 3. We assessed self-reported health-related quality of life specific to brain metastasis using the EORTC QLQ-BN20. All tests and questionnaires used were Traditional Mandarin versions and certified by a board-certified neurologist and psychologist.

Gadolinium contrast-enhanced MRI was used to assess intracranial failure and obtained prior to treatment and at 4, 9, and 12 months after WBRT until intracranial disease progression, upon new onset of neurological signs, or upon symptoms suggestive of progressive brain metastasis.

### Statistical analysis

We conducted a single-blinded randomized trial with a randomization ratio of 1:1. The actual treatment given to individual patients was determined by randomization with permuted-block design stratified by prior cranial radiosurgery. The primary endpoint was neurocognitive function as determined by a decline in HVLT-R delayed recall score from baseline to 4 months after WBRT. Previous results of standard conventional WBRT resulted in a 40% mean decline in cognitive loss at 3 to 6 months[21]. We hypothesized that using conformal WBRT with or without hippocampal avoidance would reduce the decline from baseline to 20% at 4 months. A Simon’s randomized phase II design was used to calculate sample size[23]. We required 42 evaluable subjects (21 in the HA-WBRT group and 21 in the conformal WBRT group) for a 90% probability of correctly selecting the best intervention group. We anticipated that up to 35% of patients would drop out prior to the 4-month assessment and would not be included in the final analysis. The target sample size was 64 registered subjects. Neurocognitive failure was defined as a drop in raw scores from baseline more than 2 standard deviations (SD) for any HVLT-R category (Total Recall, Delayed Recall, and Recognition Index).

Secondary endpoints included self-reported cognitive functioning, health-related quality of life, progression of brain metastasis, overall survival, and acute or late treatment related toxicity. Intracranial progression was defined as radiographic evidence of enlarged brain tumor according to RECIST 1.1 criteria or confirmed leptomeningeal seeding from cerebral spinal fluid studies. Brain progression free survival (BPFS) was calculated from randomization until brain progression or death. Hippocampal failure was documented and defined as the presence of new brain metastasis within a 5 mm margin around either hippocampus. Overall survival (OS) was defined as time from randomization to death. Self-reported cognitive functioning and health-related quality of life are under analysis and not reported in this article.

Descriptive data were reported and compared between the two arms. An independent t-test for continuous variables and Chi-square or Fisher’s exact test were performed for categorical variable comparison. A mixed effect model was used for neurocognitive function tests to assess the time effects within patients during serial follow-up and the hippocampus avoidance effect between the two arms. Survival was estimated using the Kaplan–Meier method and differences between patients or treatment characteristics were assessed using log-rank tests. A two-sided p-value of less than 0.05 was considered statistically significant. Statistical analyses were performed using GraphPad Prism 8.31 (GraphPad Software Inc.).

## Results

### Patient characteristics

From March 2015 to December 2018, we enrolled 70 eligible and evaluated 65 analyzable patients. The CONSORT diagram is shown in **Figure 1**. The median follow-up time for all evaluated patients was 12.4 months (range: 0.9 to 54.8 months). Patient characteristics are summarized in **Table 1**. There were no significant differences between the two arms in terms of age, performance status, GPA, and education level, except that more patients in the HA-WBRT arm had prior brain surgery. The representative radiotherapy plans are shown in **Supplementary Figure S1** and **Figure S2**. Despite equivalent prescription dose, patients in the HA-WBRT group received a higher integral dose compared to those in the conformal WBRT group (1158.6 MUs vs. 727.6 MUs, *p* < 0.001).

**Table 1.**
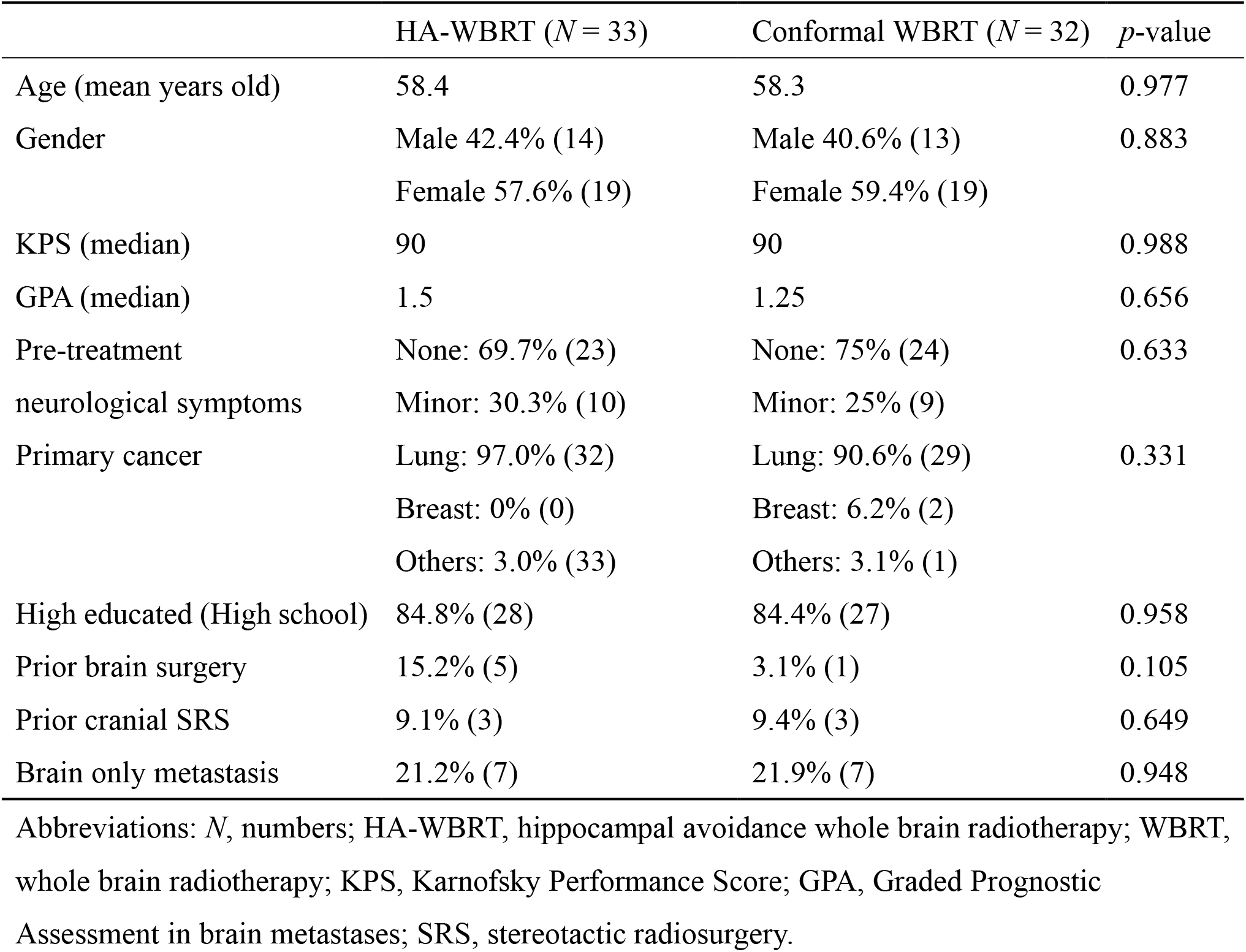
Patient characteristics

**Figure 1.**
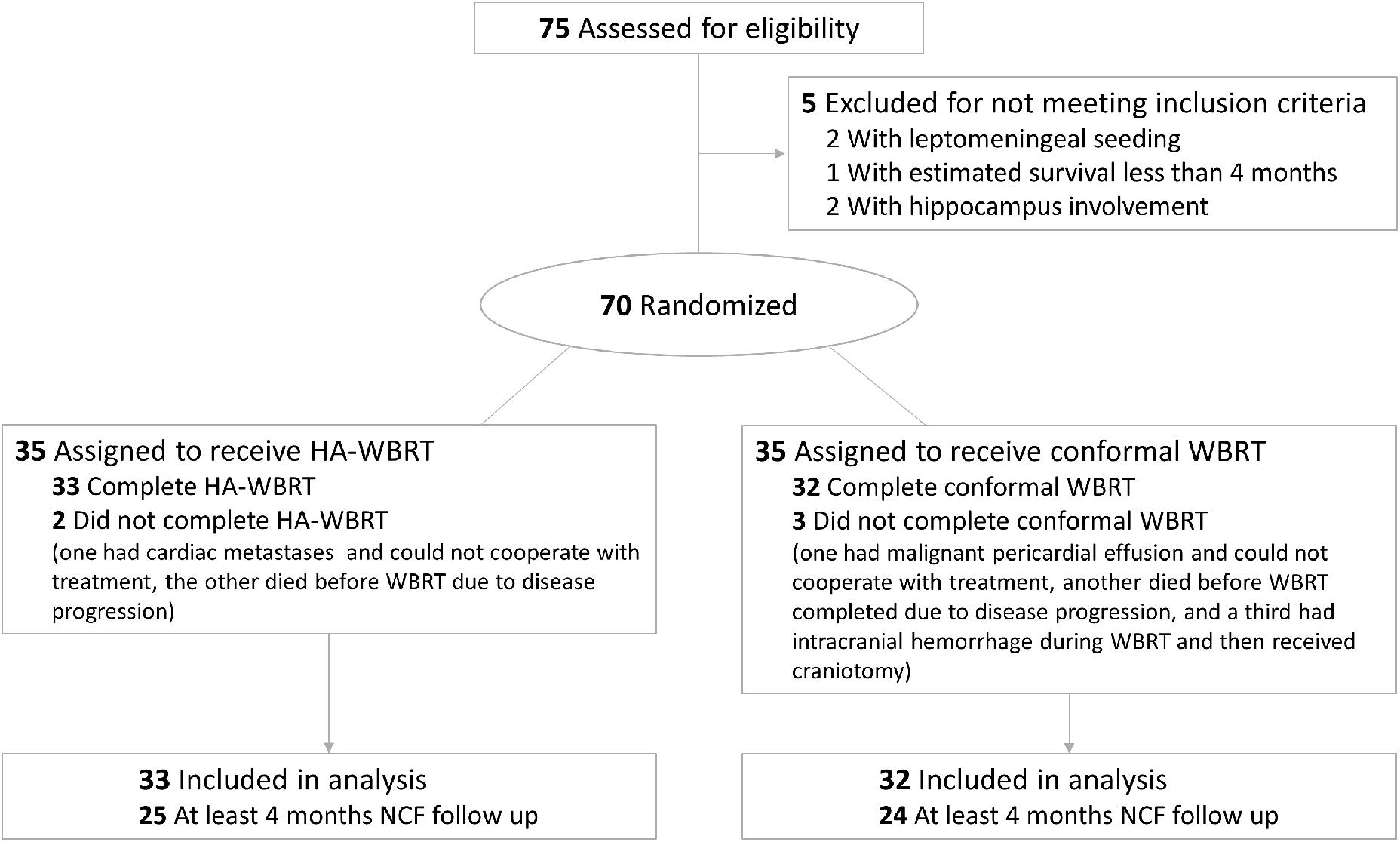
Consort flow diagram.

### Adverse events

Overall, 90.1% of patients in the HA-WBRT arm and 87.5% of patients in the conformal WBRT arm experienced some grade of toxicity. The most common toxicities were nausea (36 patients), fatigue (36 patients), vomiting (27 patients), and dizziness (9 patients). A non-significantly higher grade 2 toxicity rate was noticed in the HA-WART arm (60.6% vs. 46.9%, *p* = 0.74). One patient in each arm experienced complications with grade 3 toxicity after WBRT. One had progressive brain lesions during WBRT and received a craniotomy, while the other had severe symptomatic cerebral edema, which was resolved following bevacizumab and steroid treatment. No patients experienced grade 4 or 5 toxicity.

### Neurocognitive outcomes

All tests were performed using the Traditional Mandarin version. There were no differences in baseline neurocognitive functions between arms as shown in **Table 2**. The 4-month follow-up showed an average of –8.8% and +3.8% percentage change in HVLT-R delayed recall from baseline in the HA-WBRT arm and conformal WBRT arm, respectively. Both were better than expected and without significant differences (*p* = 0.31). Overall, there were no differences in any neurocognitive assessments between the two arms at the 4-month follow-up (**Table 2**).

**Table 2.**
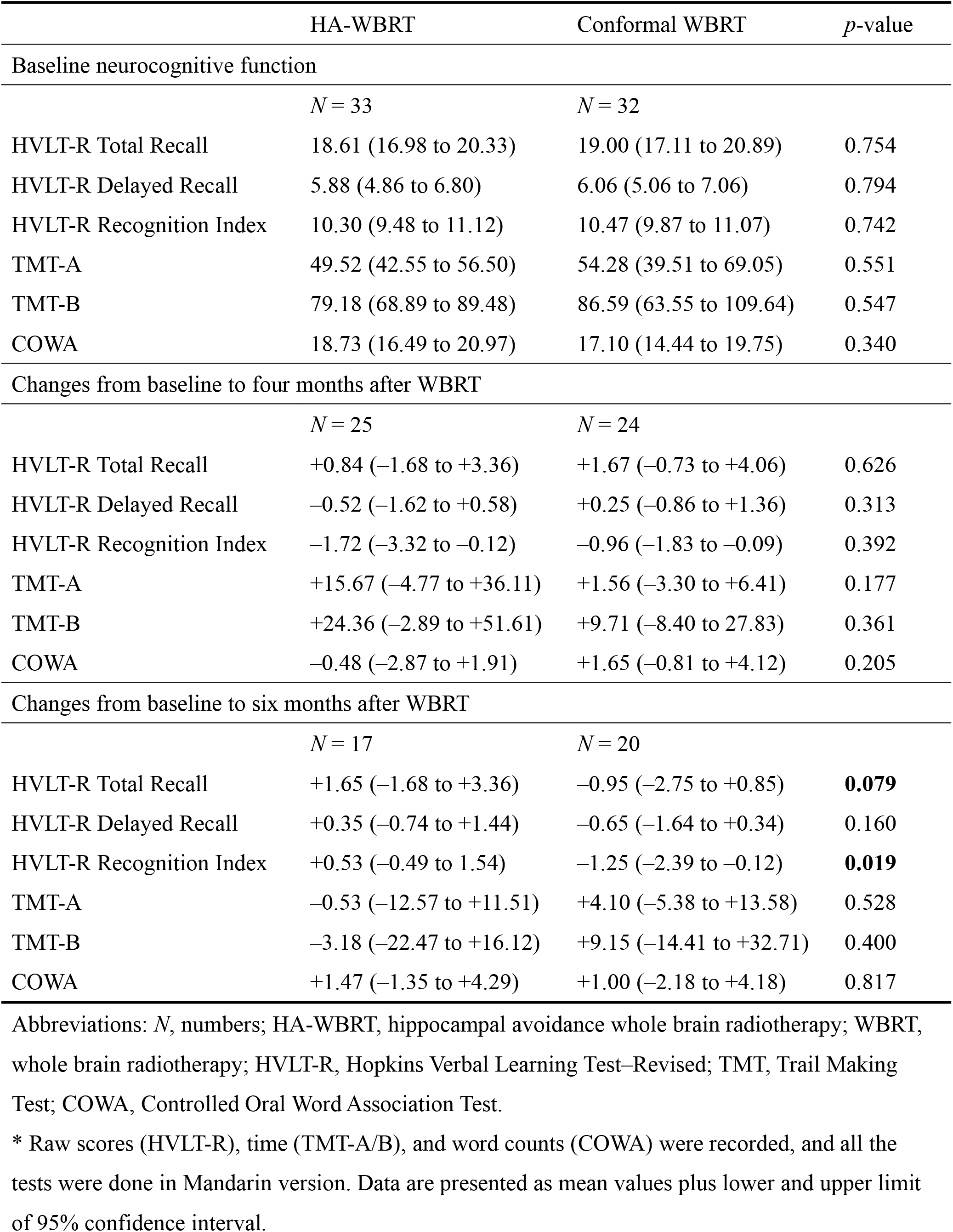
Neurocognitive function test at baseline and changes from baseline at four and six months *

|  | HA-WBRT | Conformal WBRT | *p*-value |
| --- | --- | --- | --- |
| Baseline neurocognitive function |  |  |  |
|  | *N* = 33 | *N* = 32 |  |
| HVLT-R Total Recall | 18.61 (16.98 to 20.33) | 19.00 (17.11 to 20.89) | 0.754 |
| HVLT-R Delayed Recall | 5.88 (4.86 to 6.80) | 6.06 (5.06 to 7.06) | 0.794 |
| HVLT-R Recognition Index | 10.30 (9.48 to 11.12) | 10.47 (9.87 to 11.07) | 0.742 |
| TMT-A | 49.52 (42.55 to 56.50) | 54.28 (39.51 to 69.05) | 0.551 |
| TMT-B | 79.18 (68.89 to 89.48) | 86.59 (63.55 to 109.64) | 0.547 |
| COWA | 18.73 (16.49 to 20.97) | 17.10 (14.44 to 19.75) | 0.340 |
| Changes from baseline to four months after WBRT |  |  |  |
|  | *N* = 25 | *N* = 24 |  |
| HVLT-R Total Recall | +0.84 (−1.68 to +3.36) | +1.67 (−0.73 to +4.06) | 0.626 |
| HVLT-R Delayed Recall | −0.52 (−1.62 to +0.58) | +0.25 (−0.86 to +1.36) | 0.313 |
| HVLT-R Recognition Index | −1.72 (−3.32 to −0.12) | −0.96 (−1.83 to −0.09) | 0.392 |
| TMT-A | +15.67 (−4.77 to +36.11) | +1.56 (−3.30 to +6.41) | 0.177 |
| TMT-B | +24.36 (−2.89 to +51.61) | +9.71 (−8.40 to 27.83) | 0.361 |
| COWA | −0.48 (−2.87 to +1.91) | +1.65 (−0.81 to +4.12) | 0.205 |
| Changes from baseline to six months after WBRT |  |  |  |
|  | *N* = 17 | *N* = 20 |  |
| HVLT-R Total Recall | +1.65 (−1.68 to +3.36) | −0.95 (−2.75 to +0.85) | **0.079** |
| HVLT-R Delayed Recall | +0.35 (−0.74 to +1.44) | −0.65 (−1.64 to +0.34) | 0.160 |
| HVLT-R Recognition Index | +0.53 (−0.49 to 1.54) | −1.25 (−2.39 to −0.12) | **0.019** |
| TMT-A | −0.53 (−12.57 to +11.51) | +4.10 (−5.38 to +13.58) | 0.528 |
| TMT-B | −3.18 (−22.47 to +16.12) | +9.15 (−14.41 to +32.71) | 0.400 |
| COWA | +1.47 (−1.35 to +4.29) | +1.00 (−2.18 to +4.18) | 0.817 |

Abbreviations: *N*, numbers; HA-WBRT, hippocampal avoidance whole brain radiotherapy; WBRT, whole brain radiotherapy; HVLT-R, Hopkins Verbal Learning Test–Revised; TMT, Trail Making Test; COWA, Controlled Oral Word Association Test.
\* Raw scores (HVLT-R), time (TMT-A/B), and word counts (COWA) were recorded, and all the tests were done in Mandarin version. Data are presented as mean values plus lower and upper limit of 95% confidence interval.

By analyzing all neurocognitive function tests for all time-points, we found a significant perpetuation of neurocognitive function in the HVLT-R recognition index (mean difference = 1.78, 95% confidence interval: 0.31 to 3.25, *p* = 0.019) and a trend of preservation in HVLT-R total recall (mean difference = 2.60, 95% confidence interval: –0.32 to 5.52, *p* = 0.079) at the 6-month follow-up in the HA-WBRT arm (**Figure 2** and **Table 2**). By using the memory score[24] (the sum of HVTL-R total recall and recognition index), the preservation of verbal learning and memory was significantly superior in the HA-WBRT arm at 6-month follow-up (mean difference = 4.38, 95% confidence interval: 0.72 to 8.03, *p* = 0.020), and in favor of the HA-WBRT arm thereafter till the completion of follow-up at 24 months (**Figure 2)**. Despite a lack of significant differences in HVLT-R delayed recall, TMT-A, TMT-B, or COWA tests at any time points, patients receiving HA-WBRT outperformed in all aspects of the neurocognitive function tests at the 6-month follow-up **(Table 2**). There was also no difference in cumulative incidence of neurocognitive failure between the two arms after adjusting competing risks (Gray’s test *p* = 0.93, **Supplementary Figure S3**). The 4- and 6-month cumulative incidence of neurocognitive failure was 15.2% and 18.2% in the HA-WBRT arm and 12.5% and 15.6% in the conformal WBRT arm, respectively.

**Figure 2.**
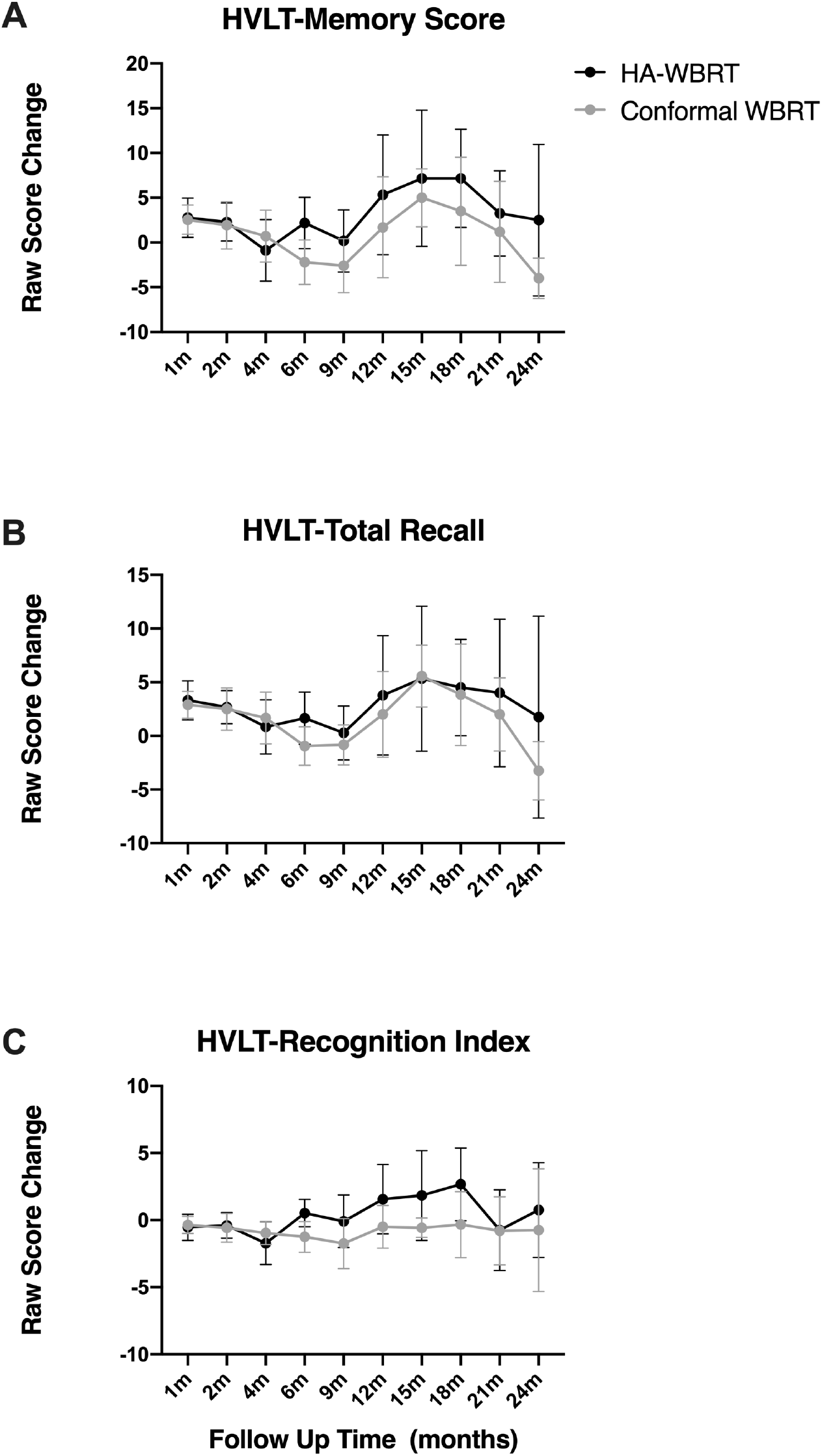
Changes in raw scores of HVLT-R (A) memory score, the sum of (B) total recall and (C) recognition Index from baseline between the hippocampal avoidance-whole brain radiotherapy (HA-WBRT) arm and the conformal WBRT arm. Data were plotted as mean values and 95% confidence intervals.

### Intracranial progression and survival

The median OS was 13.3 months for patients in the HA-WBRT arm and 15.0 months in the conformal WBRT arm (Hazard ratio = 1.32, 95% confidence interval: 0.73 to 2.38, *p* = 0.355). No significant differences in BPFS existed between the HA-WBRT and conformal WBRT arms (Hazard ratio = 1.17, 95% confidence interval: 0.69 to 1.98, *p* = 0.557). **Figure 3** shows survival curves for OS and BPFS. Adjusting the competing risk showed that there was also no difference in cumulative incidence for intracranial failure (**Supplementary Figure S4**). In total, four patients developed hippocampal failures, of which three were assigned to intervention of hippocampal avoidance (crude incidence 9.1%).

**Figure 3.**
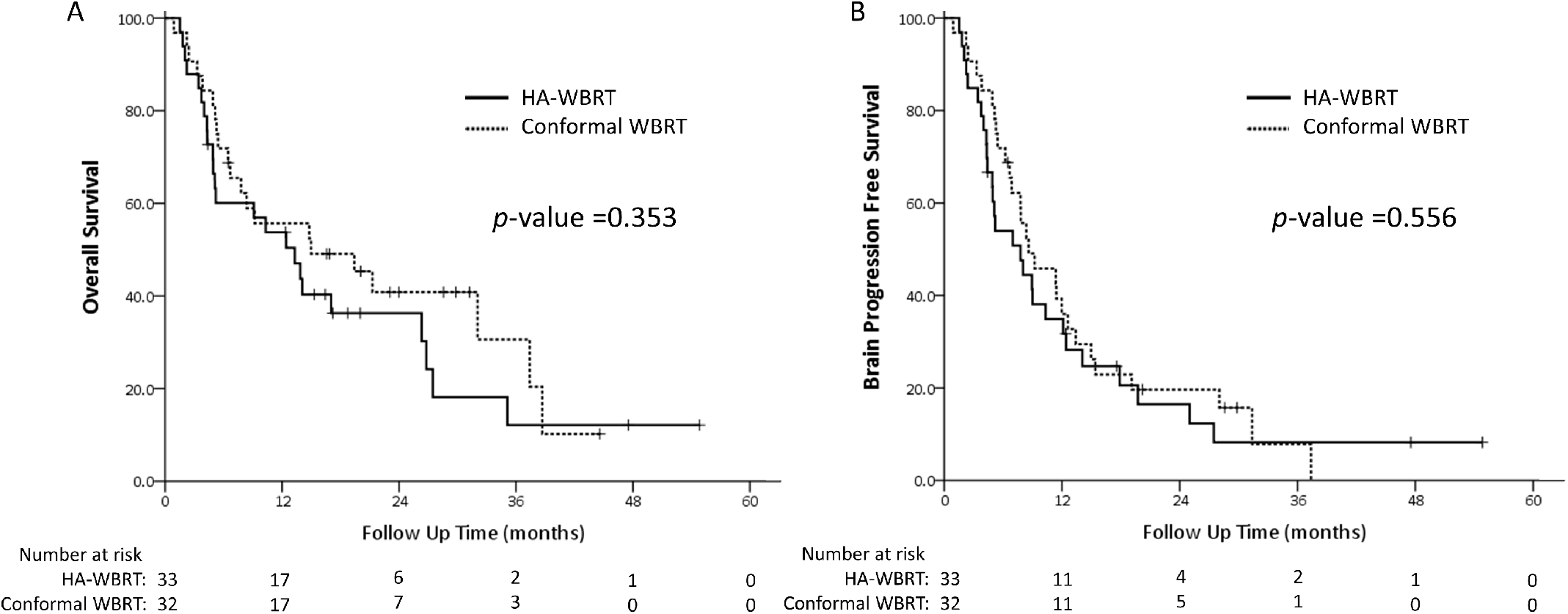
Kaplan-Meier survival curves of (A) overall survival and (B) brain progression free survival for analyzable patients between the hippocampal avoidance-whole brain radiotherapy (HA-WBRT) arm and the conformal WBRT arm.

## Discussion

This phase II randomized trial compared the effectiveness of HA-WBRT and conformal WBRT in neurocognitive function preservation. The results failed to demonstrate any benefit of hippocampal avoidance in neurocognitive preservation by HVLT-R delayed recall at four months after treatment. However, we observed a marginal benefit in perpetuation of HVLT-R total recall and significant protection in HVLT-R recognition index and memory score after HA-WBRT. To our knowledge, this is the first and only blinded randomized trial assessing the clinical benefit of hippocampal sparing performed in a non-English speaking Asian cohort. A recently published phase III randomized study NRG-CC001[25], which was a multi-center, open label trial comparing hippocampal avoidance WBRT to conventional opposing-fields WBRT together with Memantine use, demonstrated that HA-WBRT with Memantine showed significantly lower risk of neurocognitive failure (adjusted hazard ratio, 0.74; *p* = 0.02). Further, patients in the HA-WBRT arm showed less deterioration of executive function at four months (*p* = 0.01) and learning memory at six months (*p* = 0.02). In our trial, we also found less deterioration in verbal memory by HVLT-R total recall (*p* = 0.079) and HVLT-R recognition index (*p* = 0.019) at six months after WBRT. Memory score[24], which was reported to have higher sensitivity than HVLT-R total recall in detecting dementia, was also less declined in patients receiving HA-WBRT at 6 months (*p* = 0.020) and preferably preserved with longer follow-up (though non-significant). In contrast, no differences in executive function were noticed. The clinical impact of HA-WBRT seemed to be less beneficial in our study than in the NRG trial. There were some differences in trial design, which may have influenced neurocognitive outcomes.

The present study was a blinded randomized trial. Although only patients and not investigators were blinded to treatment arms, our trial could be considered a double-blind randomized trial since the neurocognitive functions were assessed independently by trained health professionals with no knowledge of the patients’ assigned group. The patient’s expectation may have some placebo or psychotherapeutic effects on neurocognitive outcomes. Open-label placebo effects are discussed extensively in the neurologic and psychiatric fields[26, 27]. Studies addressing the placebo effect on neurocognitive function have been consistently presented in various diseases, such as traumatic brain injury or Alzheimer’s disease[28, 29]. The neurobiological basis mainly involves a brain-rewarding system, where cognitive and affective functions, including awareness, insight, expectation modulation, learning, and memory, all contributed[29]. The placebo effect is also evident when treating cognitive function after radiation-induced brain injury[30].

In our comparison arm, we chose conformal WBRT rather than traditional bilateral opposing-fields WBRT. The conformal WBRT was delivered using VMAT or RapidArc techniques. The procedures for radiotherapy delivery were identical for patients in both arms, which makes the blindness more reliable. Further, conformal WBRT provided a dose distribution similar to the HA-WBRT compared with bilateral opposing-fields WBRT (**Supplementary Figure S2**). We assumed that the bias of acute toxicity to normal tissues other than the hippocampus could be minimized. The blinded testers and placebo/psychotherapeutic effects of conformal WBRT may be one reason why the difference between intervention arms was trivial in the present trial.

Memantine, a NMDA receptor antagonist, is effective in neuroprotection when in concurrent and adjuvant use with WBRT[31]. Despite its proven efficacy, the drug is not widely prescribed during WBRT in the U.S.[32]. Only 11% of radiation oncologists in the survey considered Memantine for use, with less than 10% of their patients.

Memantine was also not widely used in our society and not reimbursed by the health care system when developing the present clinical trial. Recent preclinical data revealed that Memantine may protect against radiation injury. Duman et al.[33] discovered remodeling of the hippocampal excitatory synapse following radiation treatment. Pre-administration of Memantine can revert this radiation-induced phenomenon. This interaction may imply a synergistic effect in neuroprotection from simultaneous use of HA-WBRT and Memantine. The lack of Memantine use in the present study may be another reason why patients in our cohort benefited less from HA-WBRT than those in the NRG-CC001 trial.

Importantly, current available evidence for HA-WBRT was mainly from Western countries such as the United States, especially for English-speaking cohorts. Despite these neurocognitive function tests in the neuropychiatric fields being translated into other languages, including Chinese Mandarin with validation reports[34], it is uncommon in clinical oncology trials in Asia. Effectively evaluating neurocognitive function preservation with a language barrier raises concerns. A dosimetry study of HA-WBRT showed dose volume of the irradiated hippocampus correlated well with neurocognitive function using word list learning tests in the Mandarin version[35]. They also reported no significant deterioration in memory function after HA-WBRT for patients with brain oligo-metastases or prophylactic cranial irradiation in a single arm phase II trial[36]. Our study is the first randomized trial evaluating HA-WBRT in a Mandarin-speaking cohort. The major results are in accordance with the NRG-CC001 trial, which will help reinforce the confidence of radiation oncologists in Mandarin-speaking areas to treat suitable patients with highly complex and time-consuming HA-WBRT techniques[18].

The limitations in our trial include a small patient number, unconventional phase II design, and a single institutional study. Despite those limitations, more than three quarters and half of enrolled patients followed and completed the protocol tests at 4-month and 6-month follow-ups after WBRT, respectively. The blinded testers and testees, as well as the high compliance rate of the present study, greatly reduced biases and make our results more reliable.

Combing the NRG-CC001 and present trial, both studies demonstrated that preserved verbal memory measured by HVLT-R was more prominent at 6-month rather than 4-month follow-up after HA-WBRT. This is compatible with previous clinical observations[37] and pre-clinical studies of hippocampal dysfunction[16]. Long-term follow-ups later than six months may be more appropriate to evaluate effects on verbal memory function following radiation treatment. This also suggests good-prognostic patients are more likely to benefit from HA-WBRT. Future trials should adapt late time points to determine neurocognitive outcomes.

## Conclusions

Patients receiving hippocampus-avoidant conformal WBRT without Memantine for brain metastases show better preservation of late verbal memory, but not of verbal fluency or executive function. Placebo or psychotherapeutic effects may contribute to unexpected favorable neurocognitive outcomes after conformal WBRT.

## Data Availability

Data available on request. The data underlying this article will be shared on reasonable request to the corresponding author.

## Acknowledgments

We appreciate the assistance of statistician Dr. Chin-Hao Chang, Ph.D. at the Department of Medical Research, National Taiwan University Hospital, Taipei, Taiwan, in trial design. We acknowledge the efforts of Chueh-Hui Lin, R.N. and Tsai-Tsan Wu, R.N. in trial management and data collection. We would like to thank Anthony Abram at Uni-edit for editing and proofreading this manuscript.

